# Forecasting the Diabetes Burden Across USA-Mexico Border States Through 2030: A Multi-Indicator, Multi-Model Analysis

**DOI:** 10.1101/2025.11.19.25340577

**Authors:** Kaustubh Wagh, Alexander Kirpich, Ike Okosun, Sunmi Lee, Svenn-Erik Mamelund, Gerardo Chowell

## Abstract

**Objectives:** The USA-Mexico border area faces an elevated diabetes burden, with prevalence 2-3 times higher than national averages in both countries. Yet, no forward-looking projections exist beyond 2021, limiting healthcare system planning. We provide the first multi-indicator, multi-model forecasts of diabetes burden through 2030 across border states.

**Methods:** Diabetes mortality, prevalence, and disability-adjusted life years (DALYs) data were extracted from the Global Burden of Disease Study 2021 for all ten border states in both countries (1990-2021). Eight different time-series models including ARIMA, generalized additive models, generalized linear models, Prophet, and *n*-sub-epidemic framework were used to project burden from 2022-2030. Models were evaluated and compared using weighted interval scores, with forecasts stratified by age, sex, and geography.

**Results:** Multi-model ensembles project 15-17 million people with diabetes, 23,000-26,000 annual deaths, and 1.6-1.8 million Disability-Adjusted Life Years (DALYs) due to diabetes in USA-Mexico border states by 2030. Mexican border states face accelerating mortality among adults aged 20-59 years (15-58% increases), while USA states show prevalence increases of 35-52% in youth and 47-49% in older adults. Males experienced consistently higher projected burden than females. ARIMA demonstrated superior performance for mortality and prevalence forecasting, while ensemble methods performed better for DALYs projections.

**Conclusions:** Diabetes burden is projected to increase 30-50% across border states by 2030, driven by distinct regional patterns requiring divergent public health responses. Mexican states require community health worker programs, subsidized medications, and mobile clinics for mortality prevention, while US states must expand school-based screening and nutrition program capacity to address rising youth prevalence. These projections provide actionable intelligence to support coordinated cross-border efforts in diabetes prevention and management.

**Strengths and Limitations of This Study:** *Strengths:*

– Multi-model ensemble approach (eight models) with rigorous comparative evaluation using standardized metrics, providing transparency on performance variability across outcomes and demographics.
– Comprehensive stratification by age group (five categories), sex, and geography enables demographically targeted healthcare planning, addressing a gap in prior border health forecasting studies.
– Explicit quantification of forecast uncertainty through 95% prediction intervals and formal calibration assessment, identifying outcome-specific and geographic instances of substantial uncertainty.

*Limitation:*

– Forecasts use statistically modeled disease burden estimates (GBD 2021) rather than primary surveillance data, potentially introducing measurement error and limiting precision in capturing true state-level variation.
– Time-series models assume historical trends continue unchanged, unable to account for policy interventions, technological innovations, or epidemiological shocks. Forecasts are most reliable in short term.

## 1. Introduction

The USA-Mexico border area faces a high diabetes burden, with a 2-3 times higher prevalence compared to the national average in both Mexico and the United States (1). Ten USA-Mexico border states—four in the USA (California, Arizona, New Mexico, and Texas) and six in Mexico (Baja California, Sonora, Chihuahua, Coahuila, Nuevo León, and Tamaulipas)—form a binational corridor home to over 100 million people (2,3). The border populations experience a diabetes prevalence of approximately 15.7% compared to 8.2% nationally for Mexico and 5.2% for the United States (1). With mortality rates reaching nearly 50% higher than the national averages, diabetes represents a critical and persistent public health challenge in the region (1,4,5). Border states share distinct epidemiological characteristics: higher concentrations of Hispanic/Latino populations, high poverty rates, and lower health insurance coverage compared to the national averages (6,7), alongside limited healthcare infrastructure, genetic susceptibility (e.g., TCF7L2 polymorphisms among Hispanic/Latino populations), and environmental determinants such as limited access to nutritious food, and the built environment for physical activity (8–10). This unique profile, characterized by cross-border mobility and binational health challenges, demands cross-state and country-level analysis to inform regional policy and resource allocation. Accurate and timely projections of future diabetes burden are therefore essential for state health system planning and binational coordination.

Despite extensive research on diabetes at national levels (11–13), critical gaps persist in understanding border states’ specific trajectory. To date, most diabetes forecasting studies are focused on national or global projections (14–16), with only a few examining state-level patterns in border regions. Existing forecasting studies rely on single indicators such as prevalence or mortality, limiting comprehensive burden assessment.(17,18). Moreover, few studies provide age-sex stratification for state-level interventions or compare multiple forecasting methods to quantify prediction uncertainty. While national analyses documents rising diabetes burden (17), these estimates often obscure within and between-country variation, particularly the elevated burden in border states. Our previous work documented consistently elevated historical trends in deaths, prevalence, and disability-adjusted life years (DALYs) across border states from 1990-2021, revealing consistently higher burden compared to national averages (19). However, no projections are available for these specific border states beyond 2021, leaving state policymakers without any future estimates needed to prepare healthcare systems for the coming decades. This work aims to address that and to our knowledge, this is the first state-level, multi-indicator, multi-model diabetes burden forecasting study for the USA–Mexico border region that integrates diabetes mortality, prevalence, and DALYs. This study advances prior work by comparing multiple modeling frameworks and to produce age- and sex-stratified forecasts with quantified uncertainty.

Our study employs state-level data from all ten border states (six Mexico and four USA) as the appropriate geographic unit for several reasons. First, state-level age–sex–stratified diabetes data are consistently available across decades from the Global Burden of Disease Study (20,21), ensuring comparability across countries. Second, state boundaries represent defined functional and administrative units through which health policy is formulated, healthcare systems are organized, and budgets are allocated, making state-level projections directly actionable (22). Third, the epidemiological influence of the border is broad and extends throughout border states due to population mobility, economic integration, and cultural continuity that transcends proximity to the international boundary (23,24). It is also worth emphasizing that border states exhibit distinct diabetes risk profiles compared to interior states in both countries. Fourth, state-level analysis provides sufficient data for robust age-sex stratified forecasting while enabling meaningful binational comparisons (25). Finally, sister-state relationships and regional health commissions operate at the state level, making state-level burden estimates critical for binational coordination and future collaborations. Thus, state-level analysis aims to provide optimal balance between administrative relevance, data quality, and forecasting reliability for informing border health policy.

We sought to forecast diabetes burden in all ten USA-Mexico border states from 2022 to 2030 using three complementary indicators: deaths (fatal burden), prevalence (healthcare demand), and DALYs (comprehensive disease impact combining premature mortality and disability) (26). Specific objectives include: generating projections for Mexico and USA border states to identify which region face the greatest future burden in terms of numbers; producing age-sex stratified forecasts to inform demographically targeted interventions; comparing eight time-series forecasting models (ARIMA, generalized additive models, generalized linear models, Prophet, and n-subepidemic models) to assess prediction uncertainty and identify optimal approaches for forecasting (27,28); evaluating models calibration through coverage analysis to ensure projection reliability; and comparing projected burden between U.S. and Mexican border states to inform binational coordination. These forecasts provide evidence-based estimates to support planning, binational health coordination, and targeted diabetes prevention policies.

## 2. Methods

### 2.1. Data Collection and Preparation

We obtained data for this study from the Global Burden of Disease Study 2021 (GBD2021), a publicly accessible database maintained by the Institute for Health Metrics and Evaluation (IHME) (29). We extracted diabetes-related mortality (both type 1 and type 2), prevalence, and disability-adjusted life years (DALYs) for all USA-Mexico border states from 1990 to 2021 (yearly). Border states included California, Arizona, New Mexico, and Texas on the US side, and Baja California, Sonora, Chihuahua, Coahuila, Nuevo León, and Tamaulipas on the Mexican side. We stratified all estimates by age group and sex to enable related demographic comparisons. We aggregated data for six Mexico states (referred to as MX), four USA states (referred to as US) and all ten USA-Mexico border states (referred to as USMX). The GBD 2021 data incorporate uniform and consistent methods for data collection, processing, and statistical modeling to ensure comparability across locations and time periods (30). Publicly available, de-identified aggregate data were used; institutional review board approval was not required. All data use agreements and terms of service were followed.

We used all 32 years of (yearly) historical data and set the calibration period to 32 years and the forecasting horizon between 2022 and 2030 (yearly forecasting for 9 years). This nine-year horizon aligns with the 2030 Sustainable Development Goal (SDG) targets (31), ensuring policy-relevant projections for the current global health agenda. Data was preprocessed using Microsoft Excel pivot tables. All forecasting analyses were conducted using RStudio, Posit Software, Boston, MA, USA (32) and MATLAB, MathWorks Inc., Natick, MA, USA (33). Specifically, we used the StatModPredict dashboard (developed in RStudio v. 4.5.0) to generate forecasts from the ARIMA, GAM, GLM, and Prophet models (34). For the ensemble n-sub-epidemic modeling framework, we employed the SubEpiPredict toolbox (SubEpiPredict toolbox) implemented in MATLAB (R2025) (35). Both toolboxes support reproducible workflows, automated parameter selection, and standardized output generation.

### 2.2. Forecasting Models

#### 2.2.1. ARIMA (Autoregressive Integrated Moving Average)

ARIMA is a time series model that builds forecasts by identifying patterns in past diabetes burdens (deaths, prevalence, DALYs) data (36). It combines three components: trends over time (autoregression), smoothing of random fluctuations (moving average), and differencing to stabilize the data to ensure necessary stationarity. ARIMA works the best for short-term forecasting when historical trends are stable, linear and non-seasonal (28), while it is less effective when periodic patterns shift suddenly due to interventions or emerging risk factors (28).

#### 2.2.2. GAM (Generalized Additive Model)

GAMs extend traditional regression models that allow for flexible, nonlinear relationships between time and diabetes burdens (deaths, prevalence, DALYs) (37). Instead of fitting a straight line, GAMs use smooth curves to model long-term trends in diabetes burdens (38) which make such models well suited for capturing gradual changes in diabetes burdens that may result from evolving lifestyle behaviors, treatment access, or demographic shifts.

#### 2.2.3. GLM (Generalized Linear Model)

GLMs are classic statistical models used to relate a discrete response variable (like mortality count) to one or more predictors (such as time). They assume a specific linear relationship between the predictors and the outcome, transformed by a so-called link function, and use a distribution suited for the type of data used in the analysis (e.g., Poisson or normal) (39). GLMs have straightforward formulations and easily interpretable and effective when trends are relatively smooth.

#### 2.2.4. Prophet (Developed by Meta/Facebook)

Prophet is a decomposable time series model that breaks down diabetes burdens’ (deaths, prevalence, DALYs) trends into three components: an overall trend, seasonal effects (e.g., annual patterns), and abrupt changes due to major events or interventions (40). It is available as a package in R and specifically designed to handle missing data and outliers well (41). Prophet is especially useful for longer-term forecasting where trends may shift, making it helpful for identifying inflection points in chronic disease burden.

#### 2.2.5. n-Sub-Epidemic Model

This model interprets diabetes burdens’ (deaths, prevalence, DALYs) trends as a series of overlapping “sub-epidemics”, each representing a surge or phase in diabetes deaths (42,43). Each sub-epidemic has its own timing and intensity, allowing the overall model to capture complex mortality dynamics such as plateaus or multiple peaks often seen in real-world chronic disease progression. It is particularly powerful in reflecting regional differences or the cumulative effects of repeated disruptions.

#### 2.2.6. Ensemble Sub-Epidemic Models (Unweighted and Weighted)

These models combine forecasts from the top-performing sub-epidemic models. The unweighted ensemble gives equal importance to each model, while the weighted version gives more influence to models with better fit (44). This ensemble approach improves robustness by balancing strengths across models and helps produce more reliable forecasts with quantified uncertainty. Under the n-sub-epidemic modeling framework, we developed several models, including the n-subepidemic ensemble (of top 2 ranked models) unweighted [NSE-ensemble(2)(UW)], the n-subepidemic ensemble (of top 2 ranked models) weighted [NSE-ensemble(2)(W)], and the top two individually ranked models [NSE-ranked(1) and NSE-ranked(2)]. The ensemble models [NSE-ensemble(2)(UW), and NSE-ensemble(2)(W)] are superior to individual ranked models [NSE-ranked(1), and NSE-ranked(2)] as they improve forecasting performance by systematically incorporating the predictive accuracy of individual models (44). Although the utilized n-sub-epidemic framework was initially validated primarily on infectious-disease time series, its flexible structure allows application to chronic disease burden data, where overlapping sub-trends may reflect multiple epidemiological drivers or intervention phases. However, for transparency and to show the individual models that informed our ensemble approaches, we also presented the results of NSE-ranked (1) and NSE-ranked (2) models.

### 2.3. Model Evaluation

Models were evaluated using a set of performance metrics that measure both point and probabilistic forecast accuracy simultaneously (44). In particular, the utilized models were evaluated based on Weighted Interval Score (WIS) and mean absolute error metrics, which quantify the deviation between predicted and the observed values for each given point in time and aggregated across time points. We used data from 2013 to 2021 (9 years) to measure the performance of models. In this context, smaller values of those metrics indicate better model performance. The reliability of uncertainty estimates was evaluated based on the proportion of observations falling in the predicted intervals, with 95% P.I. coverage where higher values ideally estimate better calibrated uncertainty (45). It is important to note that WIS provides a balance between sharp and out-of-range predictions due to penalizing prediction intervals that are too wide and observations that fall outside predicted range, thus sustaining an overall balance between forecast sharpness and calibration (42).

### 2.4. Quantifying Forecast

The forecasts for each model were numerically quantified. i.e. the percentage change formula was used to calculate the relative change (%) in the number of diabetes-related burdens (i.e. deaths, prevalence, DALYs) between 2021 and 2030. The exact formula was (Forecast (2030) − Observed (2021)/Observed (2021)) × 100. Here, forecast (2030) refers to the forecasted number of diabetes-related burdens in 2030 by a given model while observed (2021) refers to the observed number of diabetes-related burdens in 2021, which is the last year of available data. The overall diabetes-related burdens in 2030 were calculated by first calculating the mean forecast across all models.

Forecasts from multiple models were combined using weighted and unweighted averaging approaches. For weighted averaging, model weights were derived from the Weighted Interval Score (WIS), a proper scoring rule that evaluates both point prediction accuracy and prediction interval calibration. Weights were computed as the inverse values of WIS i.e. 1/WIS and normalized to sum to one within each geography-category stratum. Weighted forecasts were calculated by multiplying each model’s prediction by its normalized weight and summing across all models. For unweighted averaging, all models contributed equally (weight = 1/n, where n is the number of models). Prediction intervals (95% PI) were derived by aggregating the lower and upper bounds from individual models using the corresponding weighting scheme. Both methods were applied to generate forecasts separately for deaths, prevalence, and DALYs stratified by geography, sex, and age group.

## 3. Results

### 3.1 Performance Metrics

#### 3.1.1 Deaths

Mortality forecasting showed uniformly strong performance with minimal model differentiation. ARIMA, GAM, and GLM achieved comparable accuracy (MAE 2.3-3.1, WIS 2.0-3.0) across all demographic strata and geographic regions (Fig 1A, Supplementary Fig S1A). Ensemble methods showed no systematic improvement. Error magnitudes remained consistent across Mexican border states (MAE 2.5-3.0), US border states (MAE 2.8-3.3), and the combined USMX region. Model rankings were stable across age and sex strata.

**Figure 1.**
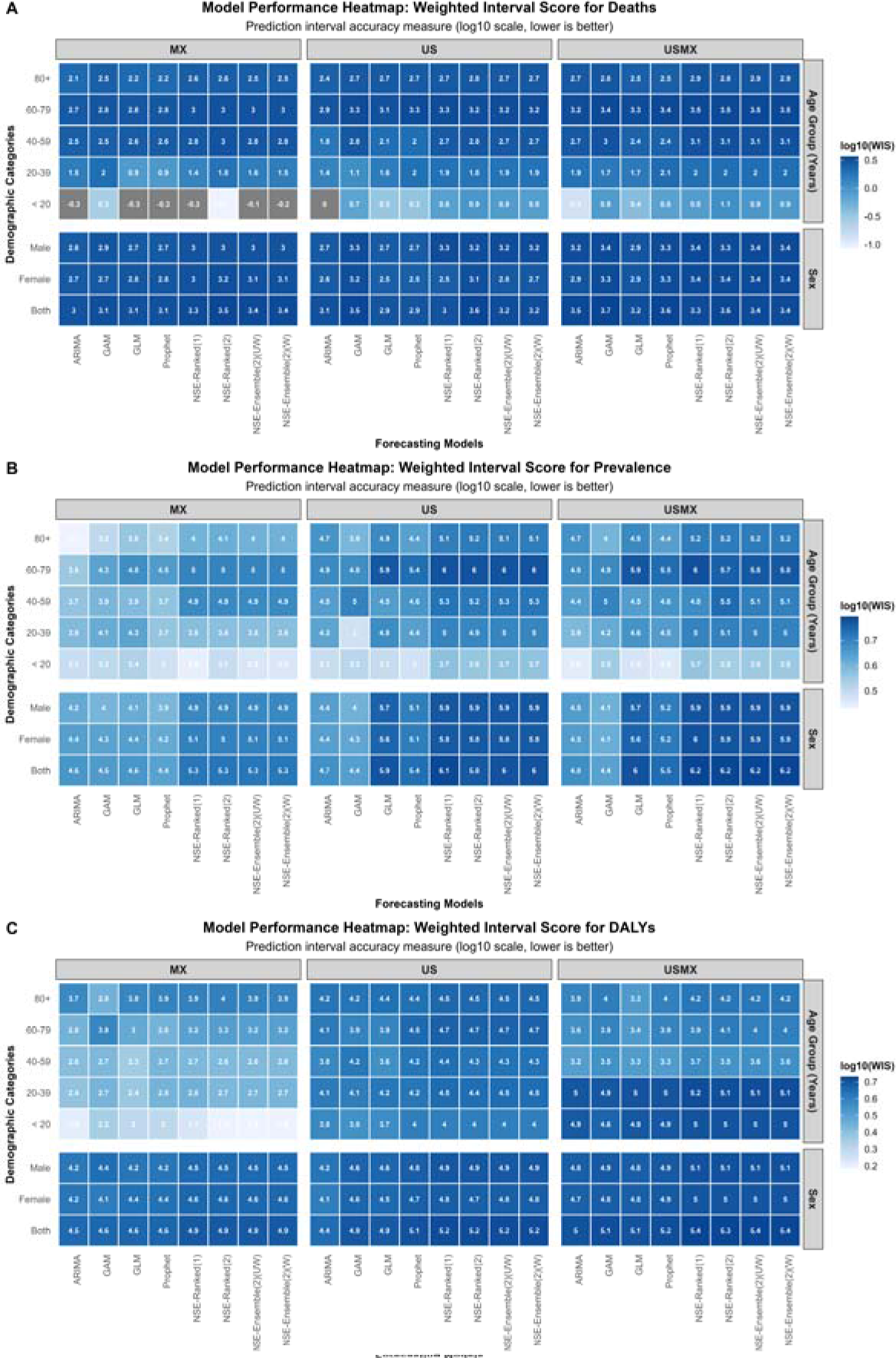
Weighted Interval Score (WIS) performance across forecasting models for diabetes burden indicators. Panel A shows deaths, Panel B shows prevalence, and Panel C shows disability-adjusted life years (DALYs). Values are displayed on a log10 scale, with lower values indicating better prediction interval accuracy. Heatmaps compare performance across geographic regions (MX, US, USMX), demographic categories (sex and age groups), and eight forecasting models including classical time series models and n-sub-epidemic (NSE) approaches.

#### 3.1.2 Prevalence

Prevalence forecasting revealed marked model performance differences among models with substantially higher errors than mortality outcomes. ARIMA achieved the lowest MAE values, ranging from 3.2-4.1 in Mexican border states to 4.5-5.0 in US border states, with WIS values following similar patterns (Fig 1B, Supplementary Fig S1B). GAM showed comparable performance. Prophet and ensemble approaches (NSE-Ensemble-Ranked, NSE-Ensemble-UW/W) consistently ranked lowest, with MAE values 15-40% higher than ARIMA across most age-sex combinations. The performance gap between ARIMA and ensemble methods was largest in Mexican border states (MAE difference 1.0-1.5 units) and smallest in US border states (MAE difference 0.5-0.8 units).

#### 3.1.3 DALYs

DALY forecasts showed error magnitudes intermediate between deaths and prevalence. ARIMA and GAM maintained lower errors (MAE 2.7-4.3, WIS 2.4-4.2) compared to Prophet and ensemble variants (MAE 3.5-5.2, WIS 3.8-5.0) (Fig 1C, Supplementary Fig S1C). The youngest age groups (<20 years) achieved MAE values 40-50% lower than older populations (60+ years) across all models. Male and female strata produced nearly identical error magnitudes and model rankings within each geographic region for all three outcomes.

### 3.2 Calibration Metrics

#### 3.2.1 Deaths

Death forecasts demonstrated the steepest age-dependent performance gradient across all outcomes (Figure 2A, Supplementary Fig S2A). Younger populations achieved lower WIS score and coverage of 95% PI closer to 95% (i.e. better calibration), with models showing WIS values of −0.7 to 0.6 log10 units and coverage of 84-100% for ages <20 years. However, GLM critically failed in middle-aged males (40-59 years), achieving only 46% coverage compared to >92% for ensemble methods. Regional disparities emerged in older populations, where Mexico consistently maintained superior coverage (>92%) across all models, while US border states required ensemble approaches to achieve adequate calibration.

**Figure 2:**
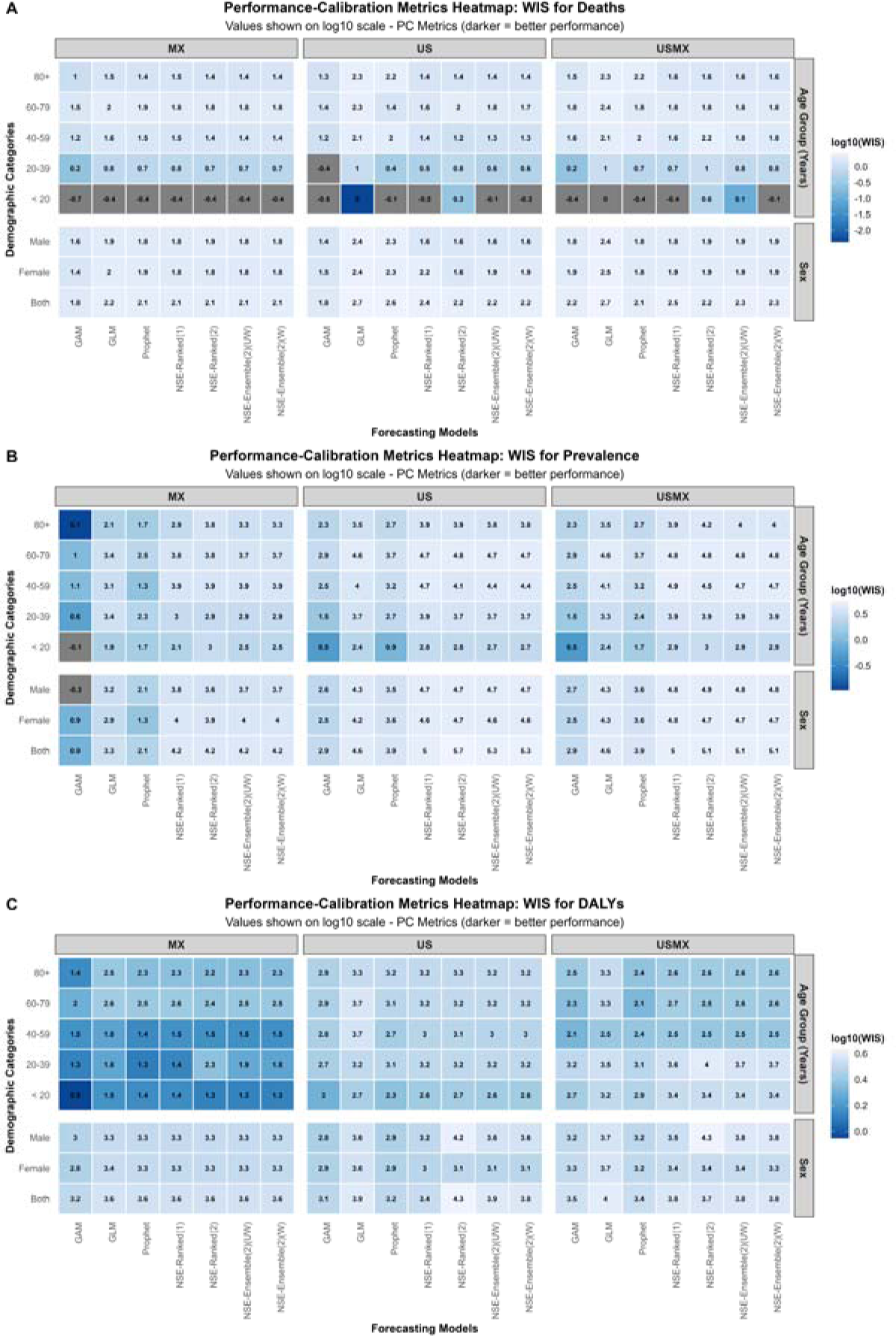
Weighted Interval Score (WIS) calibration across forecasting models for diabetes burden indicators. Panel A shows deaths, Panel B shows prevalence, and Panel C shows disability-adjusted life years (DALYs). Values are displayed on a log10 scale, with lower values indicating better prediction interval accuracy. Heatmaps compare performance across geographic regions (MX, US, USMX), demographic categories (sex and age groups), and seven forecasting models including classical time series models and n-sub-epidemic (NSE) approaches.

#### 3.2.2 Prevalence

Prevalence forecasts exhibited the widest performance variability and most severe model failures (Figure 2B, Supplementary Fig S2B). NSE-Ranked (2) catastrophically under-covered predictions in US border states for both sexes (7.69% coverage), despite acceptable performance elsewhere. This outcome showed pronounced geographic stratification: Mexico border states maintained robust calibration in younger populations (WIS: 0.5-2.9 log10 units, coverage >85%), whereas US border states struggled across all age groups, with older populations (>60 years) requiring ensemble methods to achieve minimally adequate coverage.

#### 3.2.3 DALYs

DALY forecasts revealed ensemble superiority across demographic strata (Figure 2C, Supplementary Fig S2C). NSE-Ensemble(2)(W) maintained stable performance in USMX both-sex populations (WIS: 2.5-3.3 log10 units, coverage >92%), while GAM showed age-dependent variability—excellent calibration in youth (<20 years: WIS 1.4, coverage 100%) but substantial degradation in older groups. Sex-stratified analyses showed males experienced moderately elevated WIS in young adults (20-39 years: 3.2-3.7 versus 2.8-3.4 log10 units for females). Mexico and US border states diverged primarily in middle-aged populations, where GLM coverage dropped to 46-54% in US regions.

### 3.3 Forecasting

#### 3.3.1 Deaths

Diabetes mortality forecasts from 2021-2030 varied substantially by geography (Fig 3A). MX showed the highest increases in younger adults (20-39 years: GAM 57.8%, ARIMA 14.7%) and middle-aged adults (40-59 years: GAM 40.5%, ARIMA 33.4%). US demonstrated decreases in younger populations (20-39 years: GAM −35.0%, ARIMA −10.0%) but mixed patterns in older ages. USMX showed consistent increases across all age groups (ARIMA 10-21%, GAM 10-49%). Males experienced higher mortality increases than females across all geographies (MX males: ARIMA 28.4% vs females 15.3%; US males: 21.6% vs females 3.2%). NSE models predicted mortality reductions in older US adults (60-79 years: NSE-ranked (2) −21.6%).

**Figure 3:**
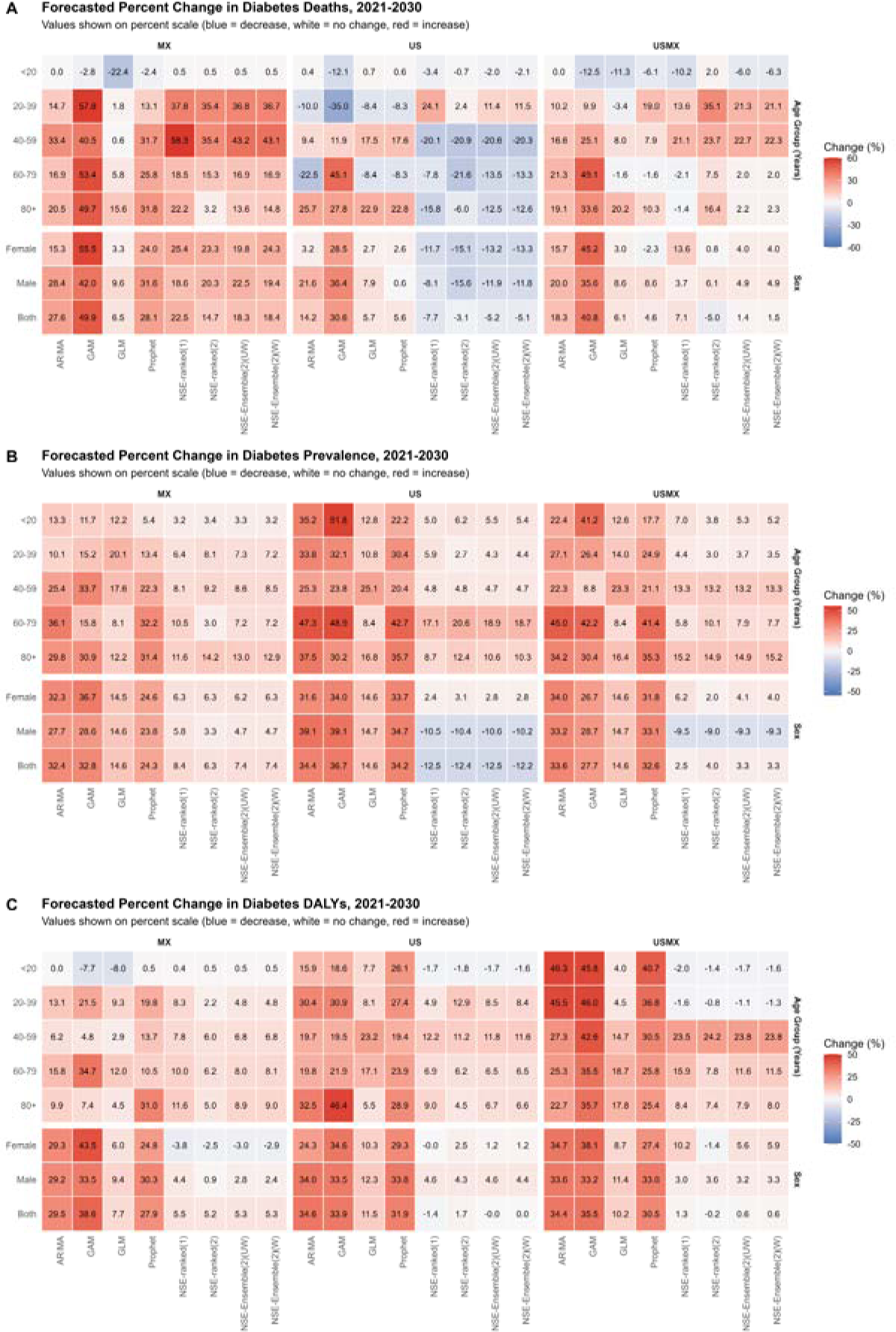
Forecasted percent change in diabetes burden indicators, 2021-2030. Panel A shows deaths, Panel B shows prevalence, and Panel C shows disability-adjusted life years (DALYs). Values represent percent change from 2021 baseline, with the color scale ranging from blue (decreases) through white (no change) to red (increases). Heatmaps compare model-based forecasts across geographic regions (MX, US, USMX), sex and age groups, and eight forecasting models including classical time series methods (ARIMA, GAM, GLM, Prophet) and neural structure ensemble (NSE) approaches.

Forecasted mortality projections varied by weighting approach (Fig 4). WIS-weighted forecasts projected 23,156 deaths (95% PI: 20,525-25,845) across the combined USA-Mexico border region, while unweighted forecasts estimated 26,419 deaths (95% PI: 22,472-30,528). Geographic distribution showed Mexico forecasted at 13,943 deaths (95% PI: 11,995-15,923) under weighted and 15,959 deaths (95% PI: 13,578-18,458) under unweighted approaches, with the United States at 12,632 (95% PI: 10,263-15,269) and 13,421 deaths (95% PI: 10,772-16,309), respectively. Males were consistently projected to experience higher mortality than females across both methods: 14,066 (95% PI: 12,556-15,631) versus 12,050 deaths (95% PI: 10,622-13,520) for weighted, and 15,368 (95% PI: 13,155-17,716) versus 14,012 deaths (95% PI: 11,896-16,253) for unweighted forecasts. Age-specific projections demonstrated strong concordance between methods, with the 60-79 age group expected to bear the highest burden under both weighted (13,012 deaths) and unweighted (14,848 deaths) approaches, followed by the 80+ and 40-59 age groups. Model-specific projected deaths by 2030 are shown in Table 1, with forecasted trajectories of deaths in Mexico and USA border states are presented in Supplementary Figs S3.1 and S3.2, respectively.

**Figure 4:**
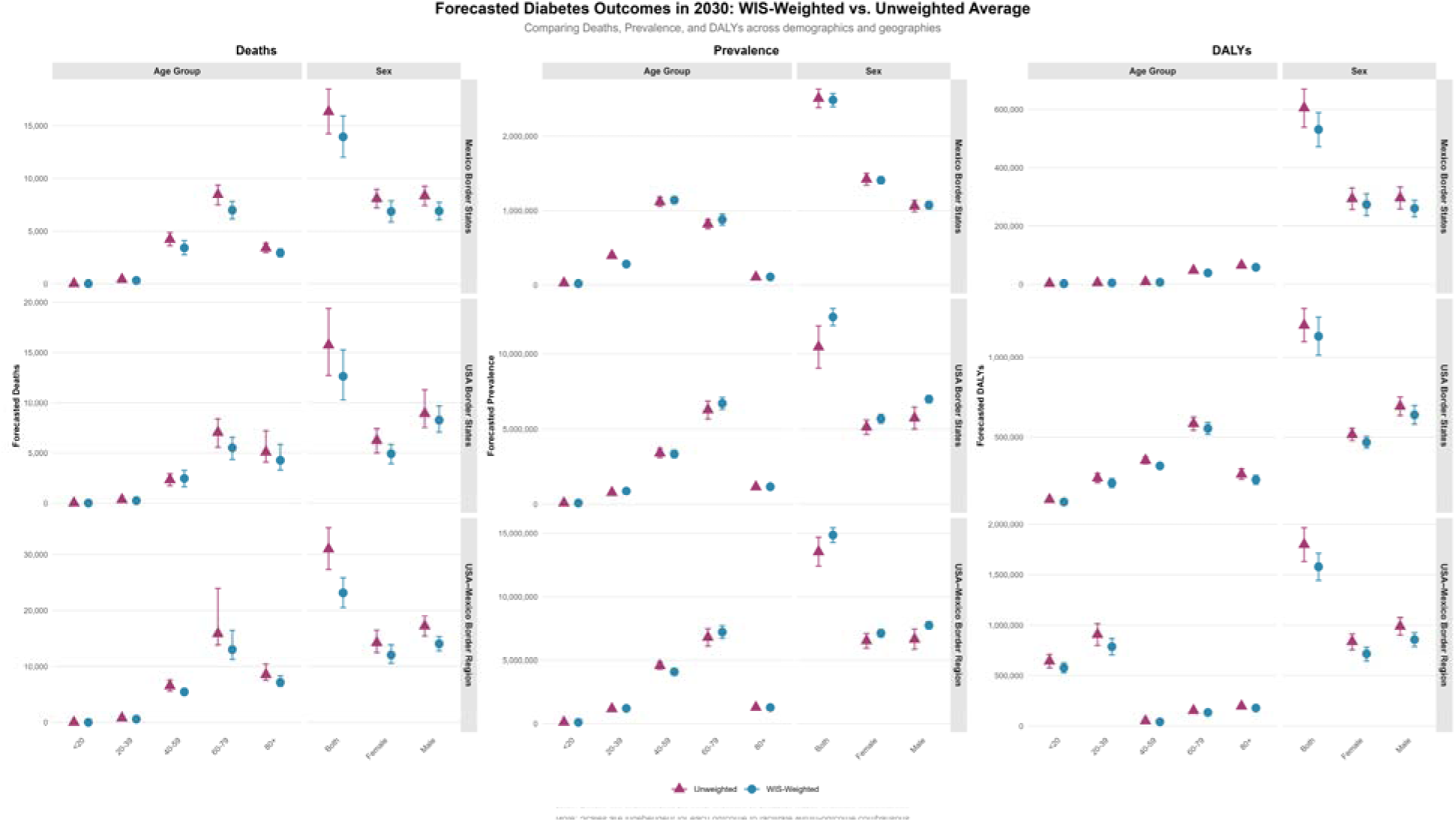
Forecasted Diabetes Burden in the USA-Mexico Border Region, 2030: Comparing WIS-Weighted and Unweighted Model Average. Point estimates and 95% prediction intervals for diabetes deaths, prevalence, and disability-adjusted life years (DALYs) in 2030. WIS-weighted averages (blue circles) assign greater weight to models with better performance compared to unweighted (pink triangles), which treats all models equally. Each outcome uses independent scales.

**Table 1.** Projected Deaths in the USA-Mexico Border Region by 2030 Stratified by Sex and Age Group. Median projected deaths (in thousands) with 95% prediction intervals across eight forecasting models. Results are presented for three geographic regions (USA-Mexico Border Region, USA Border States, Mexico Border States) stratified by sex and five age groups (<20, 20-39, 40-59, 60-79, 80+ years).

| Deaths (2030) — Median and 95% PI |  |  |  |  |  |  |  |  |
| --- | --- | --- | --- | --- | --- | --- | --- | --- |
| Category | ARIMA | GAM | GLM | Prophet | NSE-ranked(1) | NSE-ranked(2) | NSE-Ensemble(2)(UW) | NSE-Ensemble(2)(W) |
| **Sex** |  |  |  |  |  |  |  |  |
| **USA-Mexico Border Region** |  |  |  |  |  |  |  |  |
| Both | 33,523 (28,562–38,484) | 39,895 (32,365–47,425) | 30,061 (28,780–31,342) | 29,639 (27,172–31,771) | 30,348 (27,927–33,556) | 26,912 (24,263–29,630) | 28,733 (24,748–33,044) | 28,755 (24,829–33,013) |
| Female | 14,924 (12,266–17,583) | 18,723 (14,708–22,738) | 13,281 (12,556–14,005) | 12,599 (11,418–13,906) | 14,652 (12,309–16,859) | 13,001 (12,185–13,850) | 13,416 (12,214–16,489) | 13,416 (12,221–16,502) |
| Male | 18,539 (16,167–20,911) | 20,947 (17,244–24,650) | 16,780 (16,189–17,371) | 16,778 (15,657–17,887) | 16,010 (14,517–17,595) | 16,394 (14,698–17,865) | 16,205 (14,589–17,782) | 16,202 (14,580–17,759) |
| **USA Border States** |  |  |  |  |  |  |  |  |
| Both | 17,229 (11,620–22,837) | 19,708 (16,399–23,017) | 15,946 (14,467–17,426) | 15,940 (13,072–19,000) | 13,934 (11,220–19,136) | 14,617 (11,936–17,031) | 14,302 (11,508–18,318) | 14,327 (11,468–18,346) |
| Female | 6,579 (5,204–7,953) | 8,190 (6,946–9,433) | 6,544 (5,707–7,381) | 6,538 (4,849–8,091) | 5,629 (4,658–6,634) | 5,410 (4,113–6,581) | 5,535 (4,243–6,597) | 5,527 (4,295–6,584) |
| Male | 10,602 (8,381–12,843) | 11,891 (9,116–14,666) | 9,402 (8,746–10,058) | 8,771 (7,781–9,779) | 8,014 (6,898–11,486) | 7,361 (6,345–10,262) | 7,681 (6,498–10,696) | 7,688 (6,492–10,526) |
| **Mexico Border States** |  |  |  |  |  |  |  |  |
| Both | 16,903 (13,764–20,043) | 19,864 (14,259–25,469) | 14,115 (13,430–14,799) | 16,975 (16,274–17,656) | 16,228 (14,457–18,036) | 15,192 (13,792–16,479) | 15,678 (13,954–17,612) | 15,683 (13,958–17,603) |
| Female | 7,517 (6,484–8,550) | 10,139 (7,107–13,171) | 6,737 (6,392–7,082) | 8,086 (7,729–8,467) | 8,181 (7,645–8,710) | 8,042 (7,505–8,586) | 7,812 (7,201–8,489) | 8,107 (7,559–8,669) |
| Male | 8,637 (7,109–10,165) | 9,557 (6,776–12,338) | 7,378 (7,029–7,726) | 8,852 (8,513–9,174) | 7,979 (7,476–8,455) | 8,095 (7,526–8,670) | 8,243 (7,554–8,932) | 8,033 (7,490–8,584) |
| **Age groups (Years)** |  |  |  |  |  |  |  |  |
| **USA-Mexico Border Region** |  |  |  |  |  |  |  |  |
| <20 | 45 (37–53) | 39 (26–52) | 40 (38–42) | 42 (39–45) | 40 (36–45) | 46 (38–54) | 42 (37–53) | 42 (37–53) |
| 20–39 | 764 (644–883) | 762 (525–999) | 669 (636–703) | 824 (781–868) | 787 (694–903) | 936 (786–1,165) | 841 (709–1,123) | 839 (707–1,109) |
| 40–59 | 6,446 (5,450–7,443) | 6,914 (4,972–8,857) | 5,969 (5,739–6,199) | 5,966 (5,539–6,396) | 6,693 (5,789–8,211) | 6,837 (5,765–7,672) | 6,787 (5,775–7,924) | 6,763 (5,783–7,888) |
| 60–79 | 17,571 (14,328–20,814) | 21,599 (17,879–25,319) | 14,262 (13,460–15,064) | 14,262 (12,672–15,715) | 14,185 (12,792–16,229) | 15,575 (13,588–17,888) | 14,775 (12,949–16,904) | 14,772 (12,962–16,585) |
| 80+ | 9,033 (7,737–10,330) | 10,137 (8,322–11,952) | 9,121 (8,667–9,576) | 8,367 (7,875–8,871) | 7,484 (6,854–8,290) | 8,830 (7,106–11,944) | 7,759 (6,928–11,418) | 7,763 (6,935–11,096) |
| **USA Border States** |  |  |  |  |  |  |  |  |
| <20 | 22 (18–26) | 19 (11–28) | 22 (20–25) | 22 (17–27) | 21 (18–24) | 22 (19–25) | 22 (18–25) | 22 (18–25) |
| 20–39 | 324 (279–368) | 234 (126–342) | 330 (307–353) | 330 (285–374) | 447 (380–555) | 368 (316–413) | 401 (327–521) | 401 (329–525) |
| 40–59 | 2,645 (1,172–4,117) | 2,704 (1,665–3,744) | 2,839 (2,553–3,125) | 2,843 (2,253–3,372) | 1,932 (1,549–2,309) | 1,912 (1,591–2,237) | 1,919 (1,567–2,281) | 1,927 (1,590–2,267) |
| 60–79 | 5,829 (4,334–7,324) | 10,914 (9,030–12,798) | 6,888 (6,166–7,611) | 6,894 (5,438–8,262) | 6,934 (5,880–8,028) | 5,895 (4,401–7,405) | 6,505 (4,659–7,882) | 6,521 (4,660–7,892) |
| 80+ | 6,002 (3,610–8,395) | 6,102 (5,051–7,153) | 5,867 (5,252–6,482) | 5,863 (4,651–7,110) | 4,020 (3,611–4,444) | 4,486 (3,459–9,156) | 4,178 (3,562–7,518) | 4,171 (3,547–7,429) |
| **Mexico Border States** |  |  |  |  |  |  |  |  |
| <20 | 23 (16–30) | 22 (14–30) | 18 (16–20) | 22 (20–25) | 23 (21–25) | 23 (21–25) | 23 (21–25) | 23 (21–25) |
| 20–39 | 382 (319–446) | 526 (305–746) | 339 (323–355) | 377 (356–396) | 459 (414–497) | 451 (398–487) | 455 (403–494) | 455 (405–493) |
| 40–59 | 4,151 (3,184–5,117) | 4,373 (2,773–5,973) | 3,130 (2,913–3,346) | 4,098 (3,876–4,309) | 4,927 (4,393–5,276) | 4,212 (3,912–4,477) | 4,458 (3,947–5,241) | 4,453 (3,953–5,235) |
| 60–79 | 8,142 (7,193–9,092) | 10,686 (7,794–13,579) | 7,373 (7,075–7,671) | 8,762 (8,436–9,094) | 8,257 (7,467–8,991) | 8,033 (7,279–8,745) | 8,147 (7,343–8,891) | 8,148 (7,346–8,928) |
| 80+ | 3,391 (2,842–3,941) | 4,213 (3,014–5,412) | 3,255 (3,066–3,443) | 3,710 (3,533–3,894) | 3,438 (3,199–3,685) | 2,904 (2,677–3,200) | 3,197 (2,715–3,648) | 3,229 (2,711–3,647) |

Median and 95% prediction interval shown as Median (LB–UB).

#### 3.3.2 Prevalence

Universal prevalence increases characterized most forecasts, but geographic patterns varied considerably (Fig 3B). US showed the steepest rises in youth (<20 years: 35-52%) and older adults (60-79 years: 47-49%), while MX exhibited modest youth increases (11-13%) but substantial middle-aged burden (34% for 40-59 years). USMX demonstrated 22-45% increases across most ages. All geographies showed similar increases among those 80+ years (30-37%). Sex differences remained minimal across all regions, typically under 6 percentage points, with males showing marginally higher prevalence growth than females.

Prevalence forecasts showed substantial differences between weighting methodologies (Fig 4). WIS-weighted forecasts predicted 14.9 million cases (95% PI: 13.0-17.0 million) across the USA-Mexico border region, whereas unweighted forecasts projected higher prevalence at 17.4 million cases (95% PI: 14.8-20.2 million). Geographic patterns remained consistent across methods, with the United States accounting for the majority: 12.5 million cases (95% PI: 10.4-14.8 million) weighted versus 14.9 million cases (95% PI: 12.5-17.5 million) unweighted, compared to Mexico’s 2.5 million (95% PI: 2.1-2.9 million) weighted and 2.9 million (95% PI: 2.4-3.3 million) unweighted. Sex-stratified projections indicated males with slightly higher prevalence than females under both approaches: 7.8 million (95% PI: 6.7-8.9 million) versus 7.1 million (95% PI: 6.2-8.2 million) for weighted, and 9.1 million (95% PI: 7.7-10.5 million) versus 8.3 million (95% PI: 7.1-9.6 million) for unweighted forecasts. The 60-79 age group was consistently projected to demonstrate the highest disease burden across both methodologies, followed by the 40-59 age group. Model-specific projected prevalence by 2030 is shown in Table 2, with forecasted trajectories of prevalence in Mexico and USA border states are presented in Supplementary Figs S4.1 and S4.2, respectively.

**Table 2.** Projected Disease Prevalence in the USA-Mexico Border Region by 2030 Stratified by Sex and Age Group. Median projected prevalence (in thousands) with 95% prediction intervals across eight forecasting models. Results are presented for three geographic regions (USA-Mexico Border Region, USA Border States, Mexico Border States) stratified by sex and five age groups (<20, 20-39, 40-59, 60-79, 80+ years).

| Prevalence (2030) — Median and 95% PI |  |  |  |  |  |  |  |  |
| --- | --- | --- | --- | --- | --- | --- | --- | --- |
| Category | ARIMA | GAM | GLM | Prophet | NSE-ranked(1) | NSE-ranked(2) | NSE-Ensemble(2)(UW) | NSE-Ensemble(2)(W) |
| **Sex** |  |  |  |  |  |  |  |  |
| **USA-Mexico Border Region** |  |  |  |  |  |  |  |  |
| Both | 15,717 (14,399–17,036) | 15,027 (14,753–15,301) | 13,487 (13,014–13,959) | 15,598 (15,057–16,130) | 12,056 (10,502–13,560) | 12,241 (10,565–13,969) | 12,151 (10,564–13,797) | 12,157 (10,548–13,793) |
| Female | 7,577 (6,892–8,263) | 7,169 (7,036–7,302) | 6,480 (6,269–6,691) | 7,456 (7,216–7,690) | 6,005 (5,181–6,802) | 5,767 (4,959–6,575) | 5,887 (5,030–6,731) | 5,884 (5,029–6,725) |
| Male | 8,134 (7,485–8,783) | 7,860 (7,717–8,003) | 7,007 (6,745–7,269) | 8,131 (7,852–8,435) | 5,527 (4,317–6,708) | 5,557 (4,268–6,864) | 5,539 (4,297–6,752) | 5,543 (4,284–6,800) |
| **USA Border States** |  |  |  |  |  |  |  |  |
| Both | 12,927 (11,760–14,095) | 13,143 (12,941–13,344) | 11,025 (10,585–11,466) | 12,908 (12,301–13,534) | 8,417 (6,241–10,516) | 8,420 (6,212–10,725) | 8,418 (6,233–10,602) | 8,440 (6,133–10,589) |
| Female | 5,841 (5,272–6,410) | 5,945 (5,843–6,047) | 5,084 (4,889–5,279) | 5,934 (5,672–6,181) | 4,543 (3,844–5,190) | 4,577 (3,918–5,249) | 4,562 (3,882–5,223) | 4,563 (3,889–5,222) |
| Male | 7,206 (6,566–7,846) | 7,202 (7,140–7,265) | 5,941 (5,696–6,187) | 6,975 (6,677–7,273) | 4,633 (3,481–5,787) | 4,638 (3,457–5,780) | 4,631 (3,467–5,782) | 4,650 (3,492–5,767) |
| **Mexico Border States** |  |  |  |  |  |  |  |  |
| Both | 2,844 (2,622–3,067) | 2,854 (2,767–2,940) | 2,461 (2,426–2,497) | 2,670 (2,615–2,718) | 2,329 (2,181–2,475) | 2,284 (2,139–2,428) | 2,306 (2,153–2,461) | 2,307 (2,153–2,456) |
| Female | 1,613 (1,501–1,724) | 1,666 (1,617–1,715) | 1,396 (1,378–1,414) | 1,518 (1,496–1,541) | 1,295 (1,188–1,398) | 1,296 (1,190–1,407) | 1,295 (1,188–1,404) | 1,296 (1,188–1,404) |
| Male | 1,187 (1,071–1,302) | 1,195 (1,149–1,241) | 1,065 (1,047–1,084) | 1,150 (1,112–1,188) | 983 (883–1,080) | 960 (869–1,055) | 973 (875–1,071) | 973 (875–1,074) |
| **Age groups (Years)** |  |  |  |  |  |  |  |  |
| **USA-Mexico Border Region** |  |  |  |  |  |  |  |  |
| <20 | 126 (119–134) | 146 (144–147) | 116 (116–117) | 121 (116–126) | 110 (102–117) | 107 (100–114) | 109 (101–116) | 109 (101–116) |
| 20–39 | 1,320 (1,235–1,405) | 1,312 (1,300–1,325) | 1,184 (1,168–1,200) | 1,297 (1,250–1,342) | 1,084 (985–1,177) | 1,069 (969–1,168) | 1,077 (977–1,174) | 1,075 (973–1,172) |
| 40–59 | 4,827 (4,373–5,282) | 4,295 (4,136–4,454) | 4,867 (4,797–4,936) | 4,779 (4,511–5,055) | 4,471 (4,174–4,745) | 4,467 (4,098–4,849) | 4,467 (4,128–4,797) | 4,470 (4,141–4,805) |
| 60–79 | 8,156 (7,374–8,938) | 7,996 (7,761–8,230) | 6,096 (5,722–6,470) | 7,954 (7,617–8,324) | 5,952 (5,036–6,837) | 6,190 (5,228–7,097) | 6,069 (5,135–6,973) | 6,059 (5,090–6,997) |
| 80+ | 1,412 (1,268–1,557) | 1,372 (1,320–1,424) | 1,224 (1,183–1,265) | 1,424 (1,362–1,499) | 1,212 (1,071–1,351) | 1,209 (1,061–1,353) | 1,209 (1,063–1,350) | 1,212 (1,064–1,351) |
| **USA Border States** |  |  |  |  |  |  |  |  |
| <20 | 102 (95–110) | 115 (114–116) | 85 (84–87) | 93 (90–95) | 80 (74–85) | 81 (74–87) | 80 (74–86) | 80 (74–86) |
| 20–39 | 910 (836–984) | 898 (889–908) | 754 (726–782) | 886 (831–939) | 720 (638–801) | 698 (606–787) | 709 (618–796) | 710 (618–794) |
| 40–59 | 3,743 (3,278–4,209) | 3,697 (3,619–3,775) | 3,739 (3,683–3,794) | 3,597 (3,316–3,881) | 3,131 (2,749–3,507) | 3,132 (2,723–3,536) | 3,129 (2,722–3,510) | 3,128 (2,731–3,521) |
| 60–79 | 7,234 (6,481–7,987) | 7,312 (7,186–7,437) | 5,325 (4,979–5,671) | 7,011 (6,663–7,374) | 5,754 (4,944–6,530) | 5,923 (5,081–6,678) | 5,840 (4,986–6,619) | 5,829 (5,003–6,626) |
| 80+ | 1,321 (1,210–1,433) | 1,252 (1,199–1,305) | 1,123 (1,085–1,160) | 1,304 (1,244–1,376) | 1,045 (926–1,169) | 1,081 (944–1,203) | 1,064 (932–1,192) | 1,060 (935–1,191) |
| **Mexico Border States** |  |  |  |  |  |  |  |  |
| <20 | 31 (27–35) | 31 (30–31) | 31 (30–32) | 29 (25–32) | 28 (27–29) | 28 (27–29) | 28 (27–29) | 28 (27–29) |
| 20–39 | 395 (337–453) | 413 (408–418) | 430 (415–445) | 406 (377–440) | 382 (364–398) | 388 (370–405) | 385 (367–403) | 384 (367–402) |
| 40–59 | 1,202 (1,119–1,285) | 1,282 (1,212–1,351) | 1,128 (1,110–1,146) | 1,172 (1,154–1,191) | 1,036 (970–1,102) | 1,047 (963–1,128) | 1,041 (966–1,119) | 1,041 (968–1,119) |
| 60–79 | 970 (873–1,067) | 826 (808–843) | 771 (741–800) | 942 (897–991) | 788 (727–849) | 735 (660–811) | 764 (671–841) | 764 (671–841) |
| 80+ | 118 (106–129) | 119 (116–121) | 102 (98–106) | 119 (113–125) | 101 (92–111) | 104 (94–112) | 102 (93–111) | 102 (92–111) |

All values shown in thousands. Median and 95% prediction interval shown as Median (LB–UB).

#### 3.3.3 DALYs

Geographic heterogeneity in DALYs projections was pronounced (Fig 3C). USMX exhibited the highest burden increases among youth and young adults (46% for both age groups), while MX showed near-zero or negative changes in these populations. US demonstrated substantial DALYs increases in the oldest adults (80+ years: 33-46%), contrasting with lower increases in MX (7-10%) for the same age group. Middle-aged adults showed wide geographic variation, with USMX predicting 27-43% increases versus 3-6% in MX. Males experienced 4-10 percentage point greater DALYs increases than females in USMX and US.

Disability-adjusted life year projections exhibited notable variation between weighting schemes (Fig 4). WIS-weighted forecasts estimated 1.6 million DALYs (95% PI: 1.4-1.8 million) across the USA-Mexico border region, while unweighted forecasts projected 1.8 million DALYs (95% PI: 1.5-2.1 million). Geographic contributions differed by method, with the United States projected at 1.1 million DALYs (95% PI: 949,000-1.3 million) weighted versus 1.3 million (95% PI: 1.1-1.5 million) unweighted, and Mexico at 531,000 (95% PI: 451,000-617,000) weighted versus 607,000 (95% PI: 516,000-704,000) unweighted. Sex-specific projections consistently showed males accumulating higher DALYs than females: 857,000 (95% PI: 739,000-982,000) versus 716,000 (95% PI: 617,000-822,000) for weighted, and 990,000 (95% PI: 841,000-1.1 million) versus 833,000 (95% PI: 708,000-966,000) for unweighted forecasts. Both methodologies revealed similar age patterns, with peak burden projected in younger populations (20-39 and <20 age groups), reflecting substantial years of life lost, while older age groups (60-79 and 80+) showed comparatively lower DALY estimates. Model-specific projected deaths by 2030 are shown in Table 3, with forecasted trajectories of DALYs in Mexico and USA border states are presented in Supplementary Figs S5.1 and S5.2, respectively.

**Table 3.**
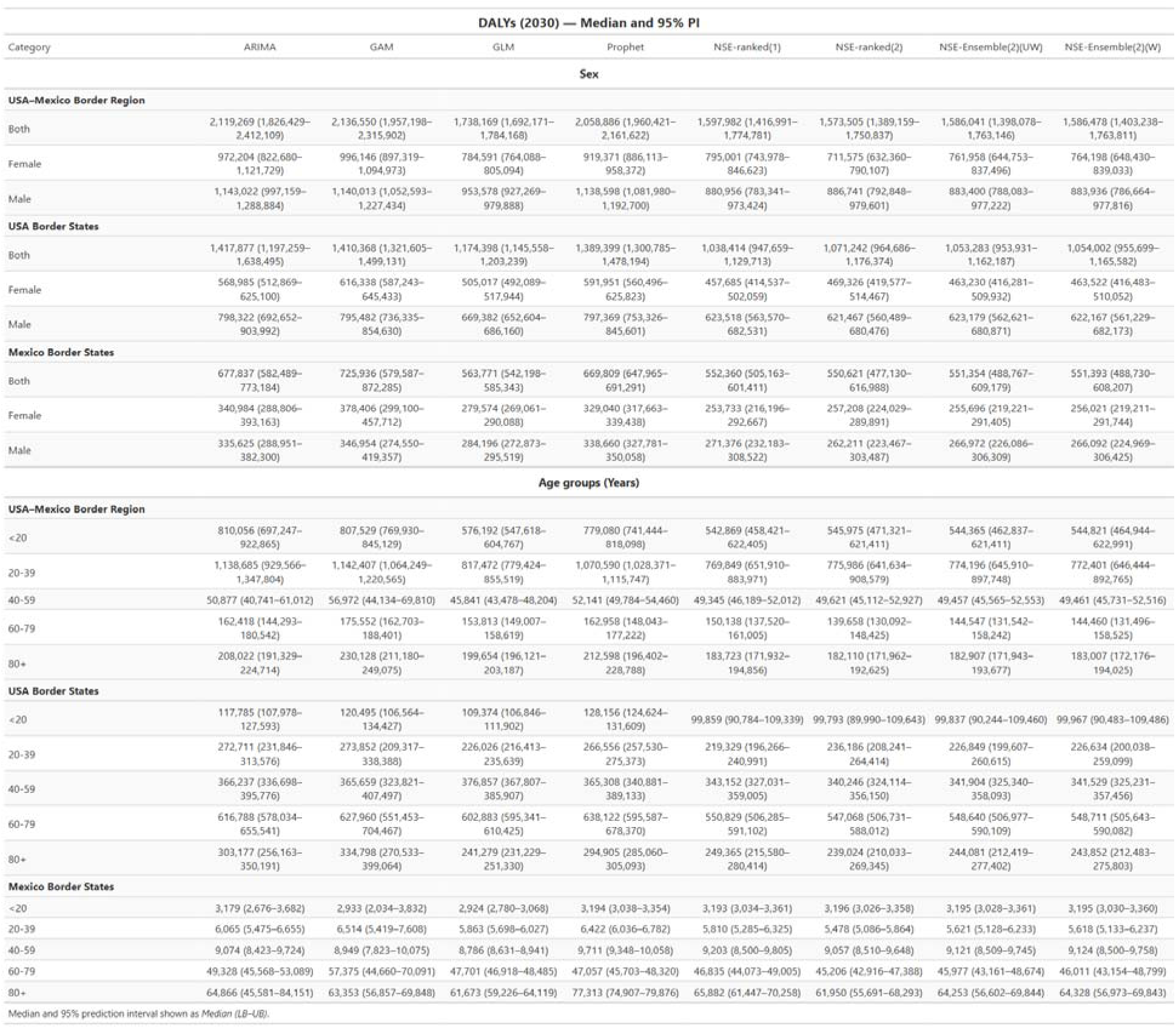
Projected Disability-Adjusted Life Years (DALYs) in the USA-Mexico Border Region by 2030 Stratified by Sex and Age Group. Median projected DALYs (in thousands) with 95% prediction intervals across eight forecasting models. Results are presented for three geographic regions (USA-Mexico Border Region, USA Border States, Mexico Border States) stratified by sex and five age groups (<20, 20-39, 40-59, 60-79, 80+ years).

Across all outcomes, ARIMA and GAM consistently produced the most reliable forecasts, demonstrating strong point prediction accuracy and similarities across demographic groups. In contrast, weighted ensemble approaches, particularly WIS-weighted models, showed much larger uncertainty in calibration and coverage performance. Together, these results highlight that no single model dominated across all indicators, emphasizing the importance of combining complementing statistical methods and model ensembles to balance accuracy and reliability in diabetes burden forecasting.

## 4. Discussion

This study provides the first comprehensive state-level forecast of the diabetes burden across all 10 USA-Mexico border states through 2030. Our multi-model ensemble approach projects approximately 23,000-26,000 diabetes deaths, 15-17 million people with diabetes, and 1.6-1.8 million DALYs annually by 2030 across the ten USA-Mexico border states, with highlighted disparities between Mexican and US border states which reflect distinct epidemiological trajectories. Overall, our findings reveal the urgent need for coordinated, binational health strategies to address the escalating diabetes burden in this region.

Our forecasts reveal three critical epidemiological patterns shaping the future diabetes burden across the border region. First, Mexican border states face accelerating diabetes mortality, particularly among younger and middle-aged adults (20-59 years), with projected increases of 15-58% by 2030. This contrasts sharply with US border states, where mortality is projected to decline in younger populations but increase among older adults. This divergence likely reflects Mexico’s ongoing epidemiological transition, characterized by rising obesity prevalence, delayed diabetes diagnosis, and limited access to glycemic control medications (12,46). The USA patterns suggests successful prevention efforts in younger cohorts but accumulated disease burden in aging populations with longer diabetes duration (47).

Second, prevalence projections uniformly indicate substantial increases across all border states, but with markedly steeper growth in US border states (35-52% in youth, 47-49% in older adults) compared to their Mexican counterpart. This pattern may reflect the expected improved survival with diabetes in the US through better treatment access, paradoxically increasing prevalence despite stabilizing incidence (48). The pronounced youth prevalence increase in US border states align with rising childhood obesity and type 2 diabetes onset at younger ages, a phenomenon extensively documented in Hispanic border communities (49,50).

Third, DALY projections highlight large increases in years of life lost among youth in USA border states, while Mexican states show stable or slightly declining DALYs in younger groups. This geographic heterogeneity with USMX projecting 46% increases in youth while Mexican states show near-zero changes suggests differential impacts of early-onset diabetes and contrasting healthcare effectiveness on different sides of the border (51).

Our findings extend and complement previous studies in several important ways. National-level projections by Lin et al. (2018) estimated US diabetes prevalence reaching 54.9 million by 2030 (11), while our border state forecasts (12.5-14.9 million) thus representing 23-27% of this expected national burden despite border states comprising only 15% of the US population, confirming the disproportionate regional impact. Similarly, Meza et al. (2015) projected Mexican national diabetes prevalence at 13.7 million by 2030 (52), with our border state expected estimates (2.5-2.9 million) representing 18-21% of the national total while border states contain approximately 20% of Mexico’s population, suggesting burden comparable to population distribution in Mexico while still elevated in absolute rate values. What is crucial is that our study addresses a few methodological gaps in previous forecasting research since most prior studies employed single models (i.e. typically ARIMA or exponential smoothing) without quantifying prediction uncertainty through detailed model comparisons (53,54). Our ensemble approach revealed substantial differences in model’s performance, particularly for prevalence forecasting where ARIMA outperformed ensemble methods by 15-40%. These finding challenges assumptions that ensemble averaging universally improves forecast accuracy (38) and suggests outcome-specific model selection strategies may be necessary and should not be discarded. The catastrophic under-coverage that was observed with certain models in US border states [7.69% for NSE-ranked (2)] underscores the importance of rigorous calibration assessment beyond point estimate accuracy. While ARIMA and GAM offered strong interpretability and transparent trend estimation, ensemble models provided greater flexibility and improved uncertainty calibration. Recognizing that the optimal model may vary by outcome type (i.e., mortality, prevalence, or DALYs) represents an important methodological contribution of this study, supporting an outcome-dependent approach to model selection.

Our age and sex stratified forecasts also address a critical limitation in existing border health research. Previous border diabetes studies provided descriptive epidemiology (55) or intervention evaluations (56) but lacked forward-looking projections with the desired level of demographic stratification details necessary for healthcare capacity planning. The pronounced sex differences identified in the analysis including males experiencing 4-10 percentage point greater increases in mortality and DALYs align with documented gender disparities in diabetes self-management, healthcare utilization, and cardiovascular complications among Hispanic populations (51).

Since GBD 2023 data were released after our initial analysis, we compared our projections with these independent estimates as a validation check. Our multi-model ensemble forecast 11.6 million (95% PI: 10.7–12.5 million) people with diabetes in the USA-Mexico Border (USMX) region for 2023, which closely aligned with the GBD estimate of 11.4 million (95% uncertainty interval: 10.6–12.3 million). Similarly, our forecast of 13–15 thousand diabetes-related deaths in six Mexico border states (MX) in 2023 was consistent with the GBD estimate of 13–16 thousand deaths.

### 4.1 Public Health and Healthcare System Implications

The diabetes projections for the USA-Mexico border highlighted in this study reveal a pressing public health issue that requires urgent attention. First, it is essential to develop a coordinated binational health policy and allocate resources effectively. Prioritizing initiatives that promote diabetes health by supporting weight management, encouraging regular physical activity, and promoting a balanced diet rich in fiber, fruits, vegetables, and whole grains are warranted.

Second, Mexican border states require immediate strengthening of primary care infrastructure for diabetes screening and early treatment. The projected mortality increase among working-age adults (20-59 years) is expected to generate substantial economic losses through premature death and disability (57). Targeted interventions should include community health worker programs, which have demonstrated effectiveness in Hispanic border communities (58), and improved access to metformin, statins, and antihypertensive medications through public pharmacy networks. Third, US border states must expand healthcare capacity to be ready to accommodate 35-52% prevalence increases in youth populations. This requires school-based screening programs, pediatric endocrinology workforce expansion, and integration of behavioral health services to address psychosocial barriers to diabetes self-management in Hispanic youth (59). The projected burden which is concentrated in the 60-79 age group necessitates geriatric care infrastructure investments and age-appropriate disease management programs. Four, binational coordination mechanisms, including the USA-Mexico Border Health Commission and sister-state partnerships, should consider utilizing these state-specific projections to align policy priorities and share evidence-based interventions. In particular, the divergent epidemiological trajectories we identified suggest opportunities for bidirectional learning, with Mexico potentially adopting similar policies and benefiting from US prevention successes in younger populations while the US could adopt Mexico’s community-based care models that maintain a lower DALY burden in older adults despite resource constraints (55). By taking the above actions, we can significantly enhance diabetes health and outcomes for communities on both sides of the border.

### 4.2 Limitations

Several limitations warrant consideration. First, forecasts assume historical trends continue without accounting for potential policy interventions, technological innovations (e.g. improved medications and treatment), or epidemiological shocks that could alter trajectories (60). Second, the utilized GBD estimates rely on statistical modeling rather than comprehensive surveillance systems in some border states, potentially introducing measurement error (26). Third, the GBD model estimates themselves are not derived from primary surveillance data at the state level, which may limit precision in capturing substate variations. Fourth, our models do not incorporate diabetes type differentiation (i.e. Type 1,and Type 2), fertility and migration patterns, or cross-border healthcare utilization factors affects border regions (61). Fourth, forecasting performance varied substantially by outcome (data type) and geography (state), with prevalence projections in US border states showing wider prediction intervals than mortality forecasts. Finally, our study period ended in 2021, precluding assessment of potential pandemic-related disruptions to diabetes care and screening that may influence future burden (62).

## 5. Conclusion

Diabetes burden across USA-Mexico border states will increase 30-50% by 2030, with divergent epidemiological trajectories requiring distinct binational responses. Mexican border states demand immediate mortality prevention through expanded treatment access, while US border states must prepare healthcare infrastructure for increases in youth prevalence. These evidence-based projections provide actionable intelligence for resource allocation, healthcare system strengthening, and binational policy coordination to mitigate the escalating diabetes crisis throughout the joint border region. Beyond diabetes, the utilized forecasting framework can be applied to other chronic diseases and cross-border settings to support evidence-based planning and progress toward global health targets.

## Data Availability

The datasets generated and analyzed during the current study are available publicly as described in the methods section.

https://vizhub.healthdata.org/gbd-results/

## Acknowledgments

The authors would like to thank Hamid Karami, Georgia State University, for sharing his valuable expertise in MATLAB coding through hemera.rs.gsu.edu (interactive online entry point to advanced research computing) which greatly contributed to the success of this research.

## Author Contributions

KW: design, analysis, interpretation, drafting, and editing; AK: review, and editing; IO: review, and editing; SL: review; SEM: review, and editing; GC: conception, supervision, review, and editing.

