## Supplementary Figures for "Forecasting the Diabetes Burden Across USA-Mexico Border States Through 2030: A Multi-Indicator, Multi-Model Analysis"

### Supplementary Figures and Tables

#### Supplementary Fig S1. Mean Absolute Error (MAE) performance across forecasting models for diabetes burden indicators.

Panel A shows deaths, Panel B shows prevalence, and Panel C shows disability-adjusted life years (DALYs). Values are displayed on a log10 scale, with lower values indicating better prediction interval accuracy. Heatmaps compare performance across geographic regions (MX, US, USMX), demographic categories (sex and age groups), and eight forecasting models including classical time series models and n-sub-epidemic (NSE) approaches.

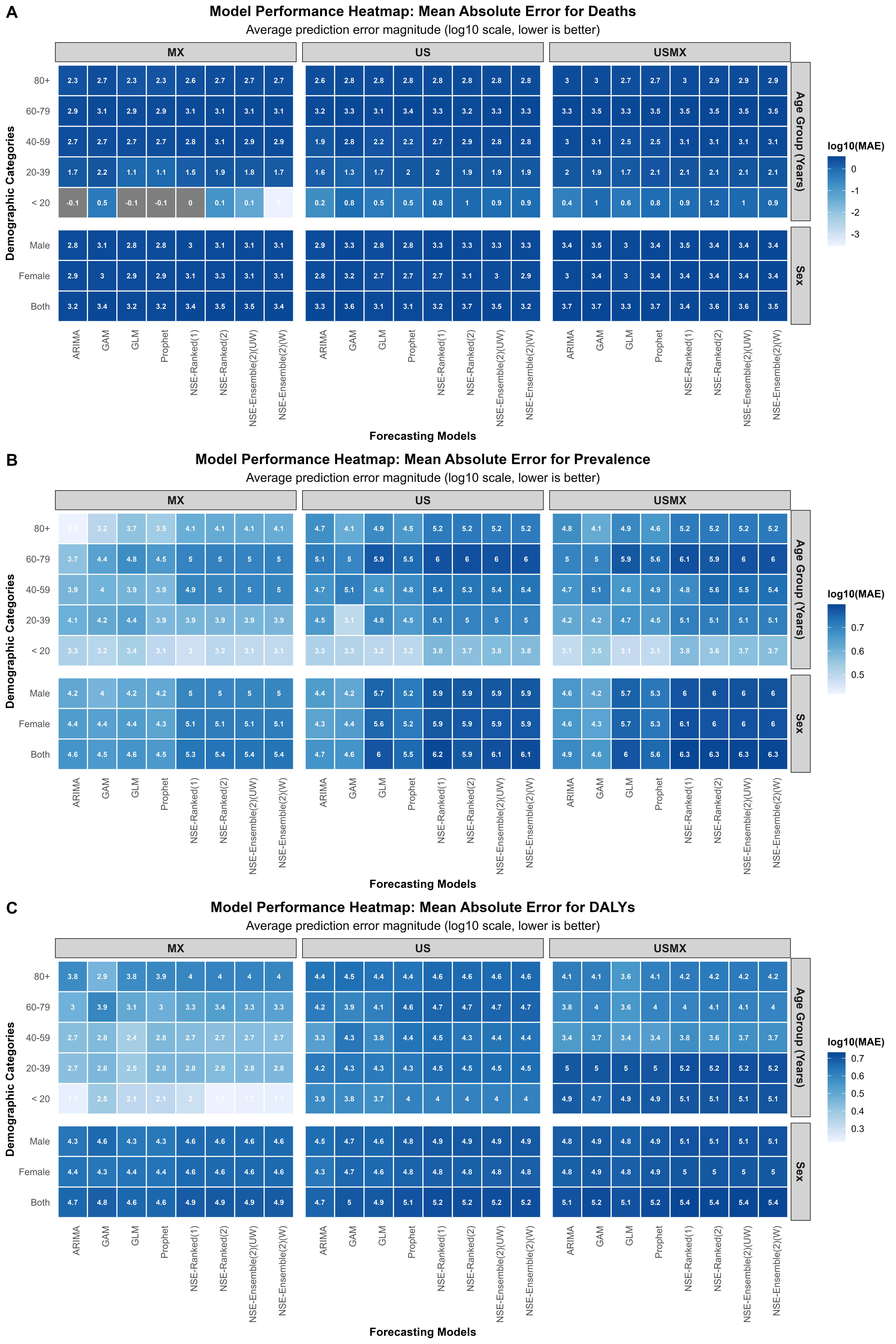

#### Supplementary Fig S2. Coverage 95% Prediction Interval calibration across forecasting models for diabetes burden indicators.

Panel A shows deaths, Panel B shows prevalence, and Panel C shows disability-adjusted life years (DALYs). Values are displayed on a log10 scale, with lower values indicating better prediction interval accuracy. Heatmaps compare performance across geographic regions (MX, US, USMX), demographic categories (sex and age groups), and seven forecasting models including classical time series models and n-sub-epidemic (NSE) approaches.

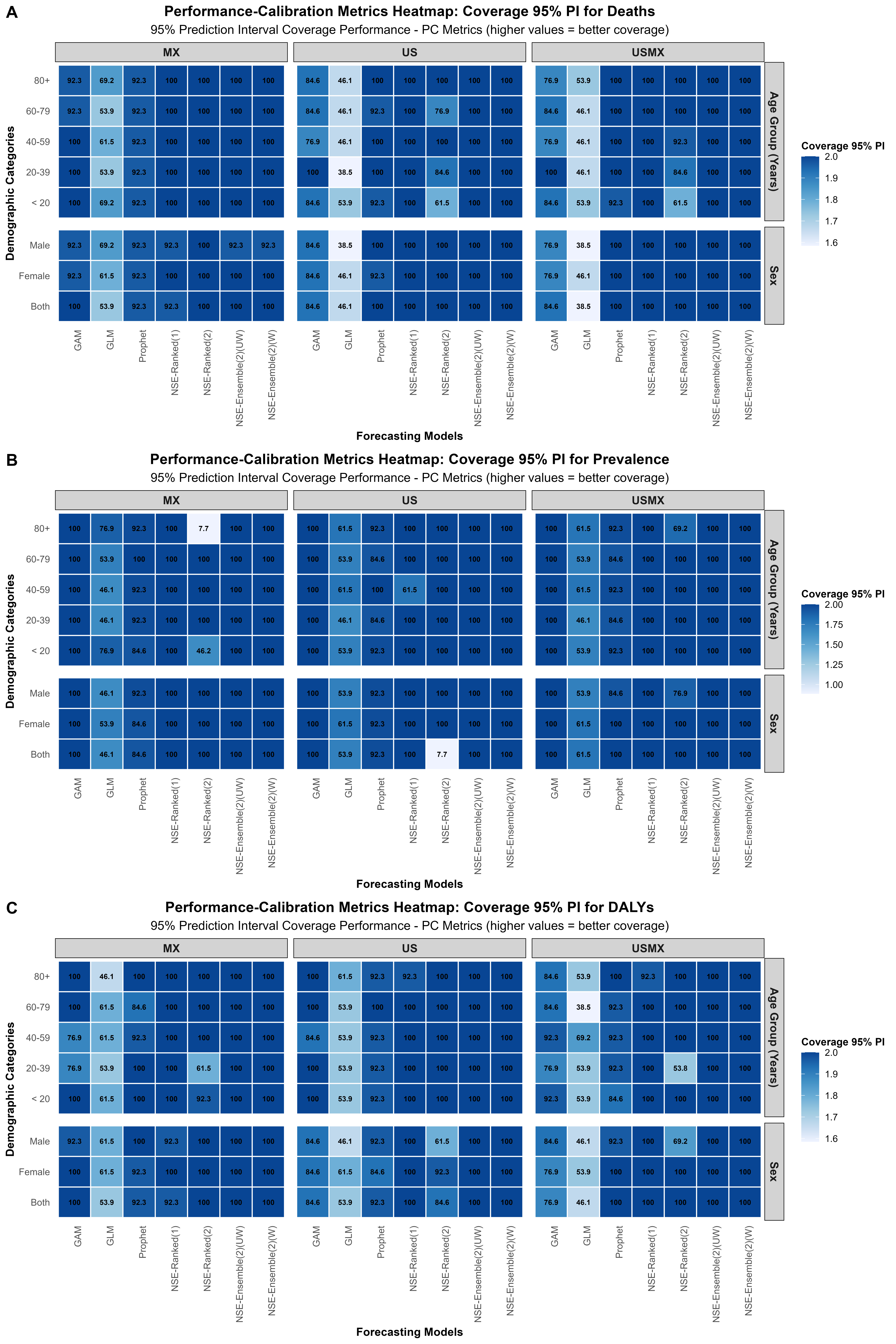

#### Supplementary Fig S3.1 Diabetes Deaths forecasts by demographic groups in Mexico, 1990-2030.

Model comparison showing historical observations (circles) and forecasts (red lines) with 95% prediction intervals (dotted lines) for diabetes-related disability-adjusted life years across eight forecasting models (ARIMA, GAM, GLM, Prophet, NSE-Ensemble variations, and NSE-Ranked variations). Data are stratified by sex (both sexes, female, male) and age groups (<20, 20-39, 40-59, 60-79, 80+ years). The vertical dashed line indicates forecast period start (2021).

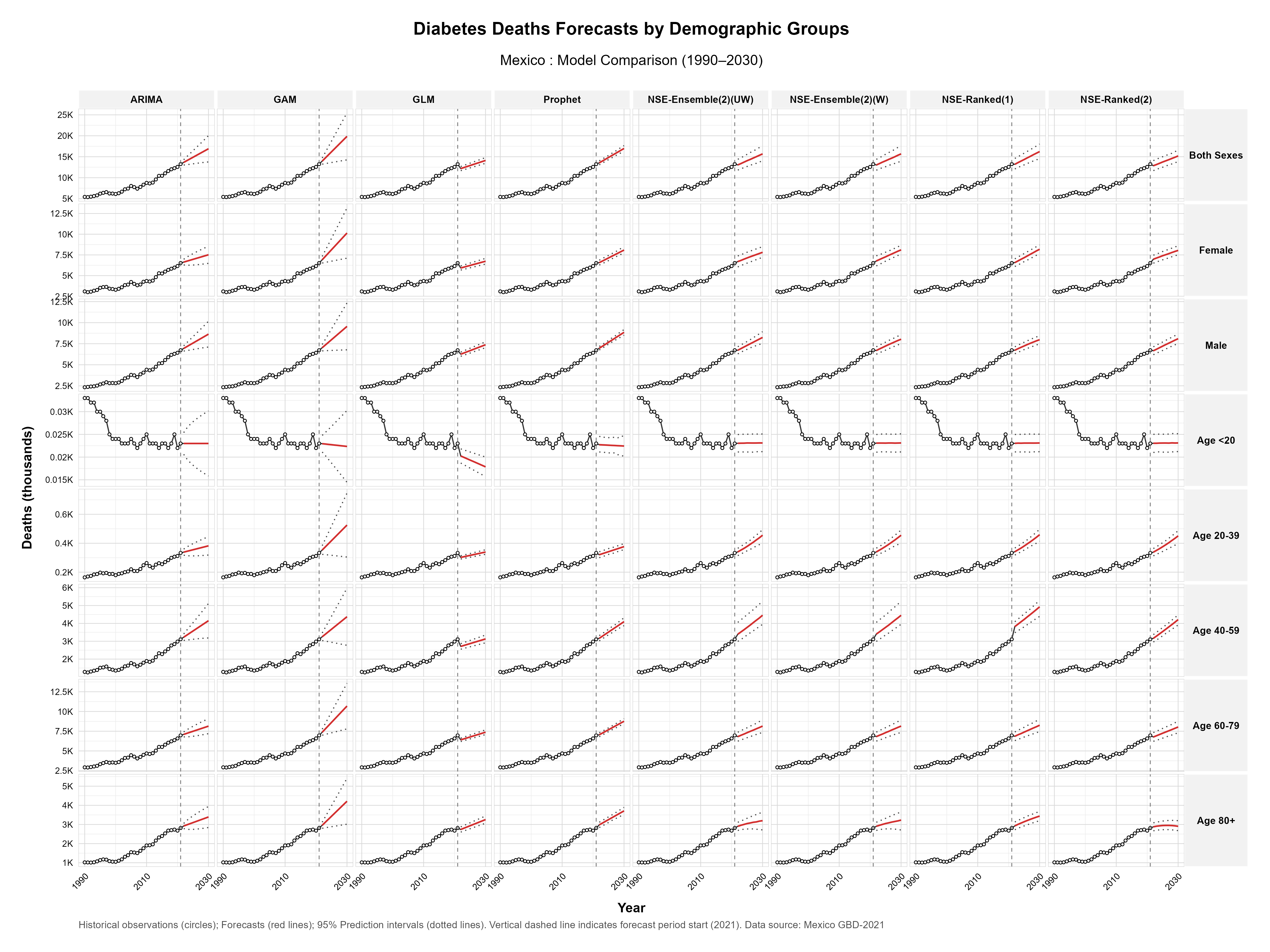

#### Supplementary Fig S3.2 Diabetes Deaths forecasts by demographic groups in USA, 1990-2030.

Model comparison showing historical observations (circles) and forecasts (red lines) with 95% prediction intervals (dotted lines) for diabetes-related disability-adjusted life years across eight forecasting models (ARIMA, GAM, GLM, Prophet, NSE-Ensemble variations, and NSE-Ranked variations). Data are stratified by sex (both sexes, female, male) and age groups (<20, 20-39, 40-59, 60-79, 80+ years). The vertical dashed line indicates forecast period start (2021).

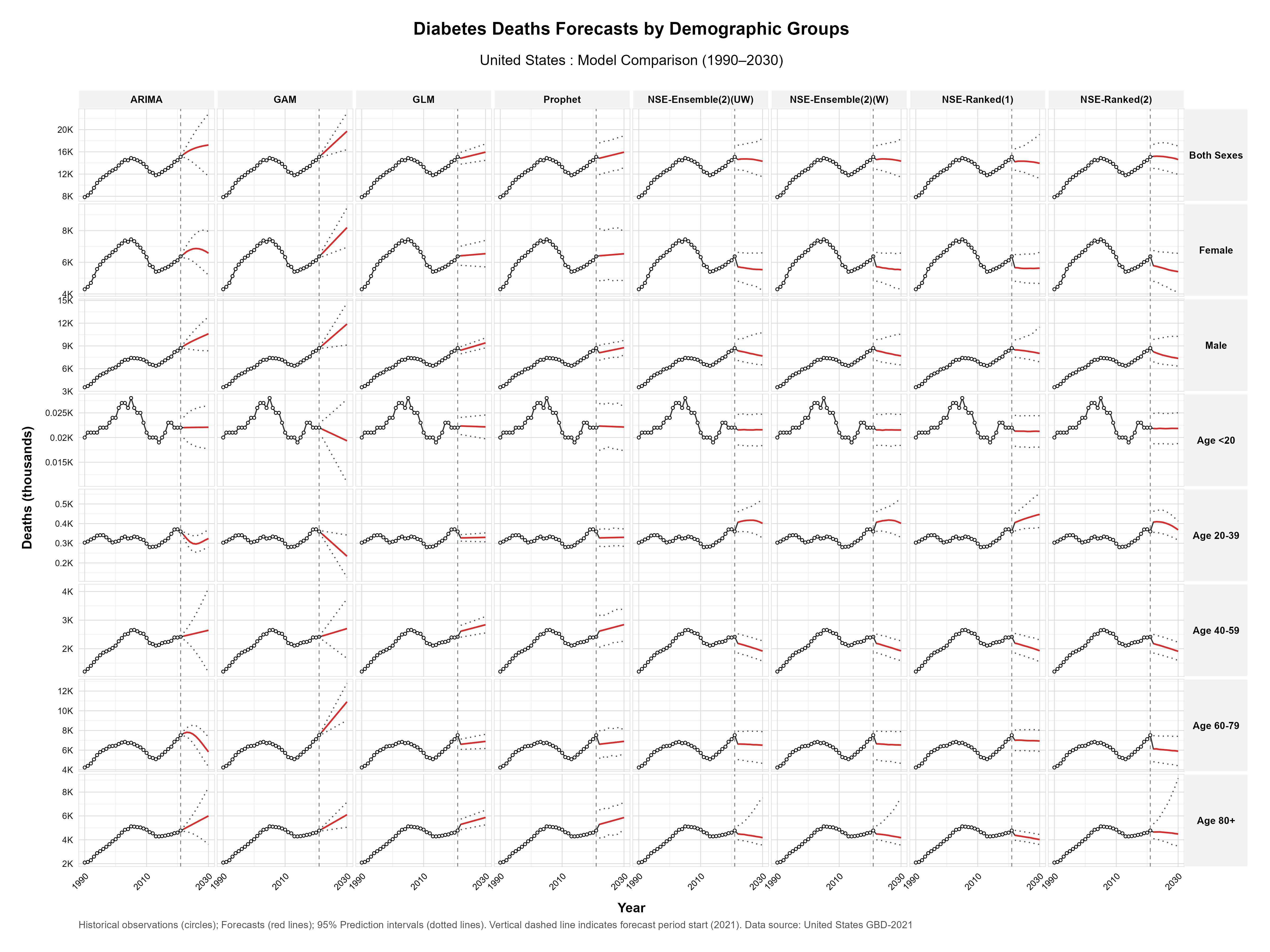

#### Supplementary Fig S4.1 Diabetes Prevalence forecasts by demographic groups in Mexico, 1990-2030.

Model comparison showing historical observations (circles) and forecasts (red lines) with 95% prediction intervals (dotted lines) for diabetes-related disability-adjusted life years across eight forecasting models (ARIMA, GAM, GLM, Prophet, NSE-Ensemble variations, and NSE-Ranked variations). Data are stratified by sex (both sexes, female, male) and age groups (<20, 20-39, 40-59, 60-79, 80+ years). The vertical dashed line indicates forecast period start (2021).

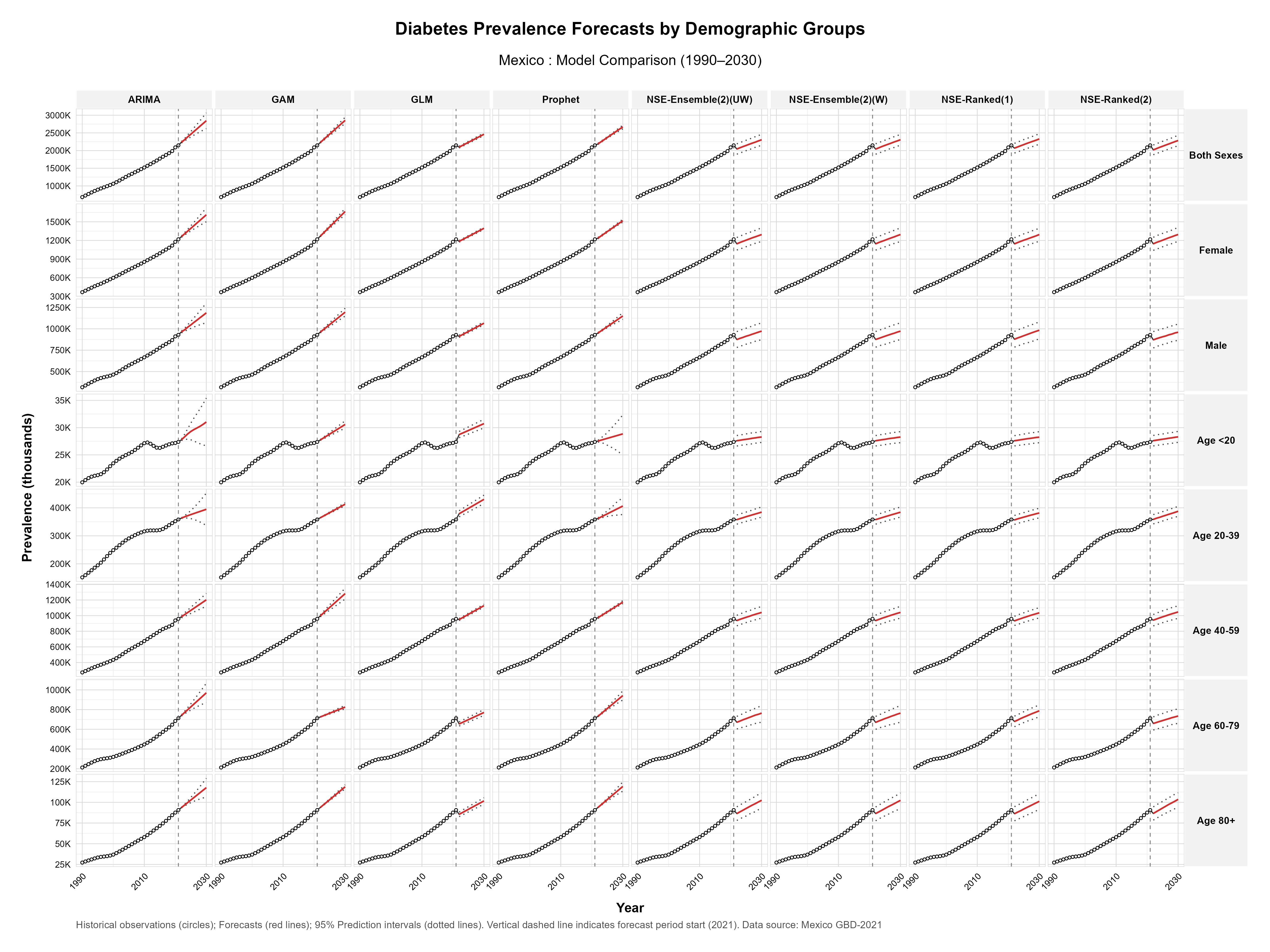

#### Supplementary Fig S4.2 Diabetes Prevalence forecasts by demographic groups in USA, 1990-2030.

Model comparison showing historical observations (circles) and forecasts (red lines) with 95% prediction intervals (dotted lines) for diabetes-related disability-adjusted life years across eight forecasting models (ARIMA, GAM, GLM, Prophet, NSE-Ensemble variations, and NSE-Ranked variations). Data are stratified by sex (both sexes, female, male) and age groups (<20, 20-39, 40-59, 60-79, 80+ years). The vertical dashed line indicates forecast period start (2021).

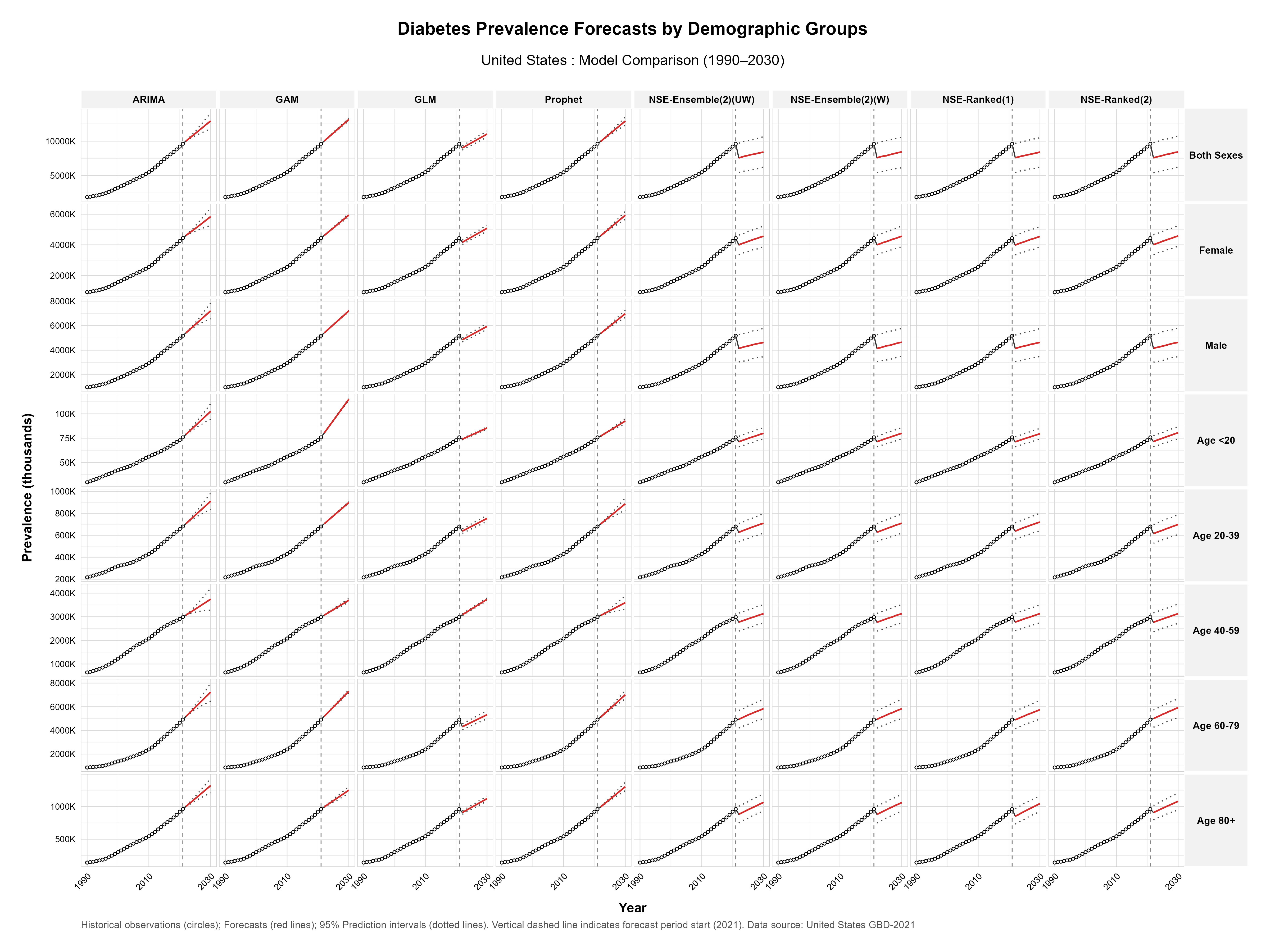

#### Supplementary Fig S5.1 Diabetes DALYs forecasts by demographic groups in Mexico, 1990-2030.

Model comparison showing historical observations (circles) and forecasts (red lines) with 95% prediction intervals (dotted lines) for diabetes-related disability-adjusted life years across eight forecasting models (ARIMA, GAM, GLM, Prophet, NSE-Ensemble variations, and NSE-Ranked variations). Data are stratified by sex (both sexes, female, male) and age groups (<20, 20-39, 40-59, 60-79, 80+ years). The vertical dashed line indicates forecast period start (2021).

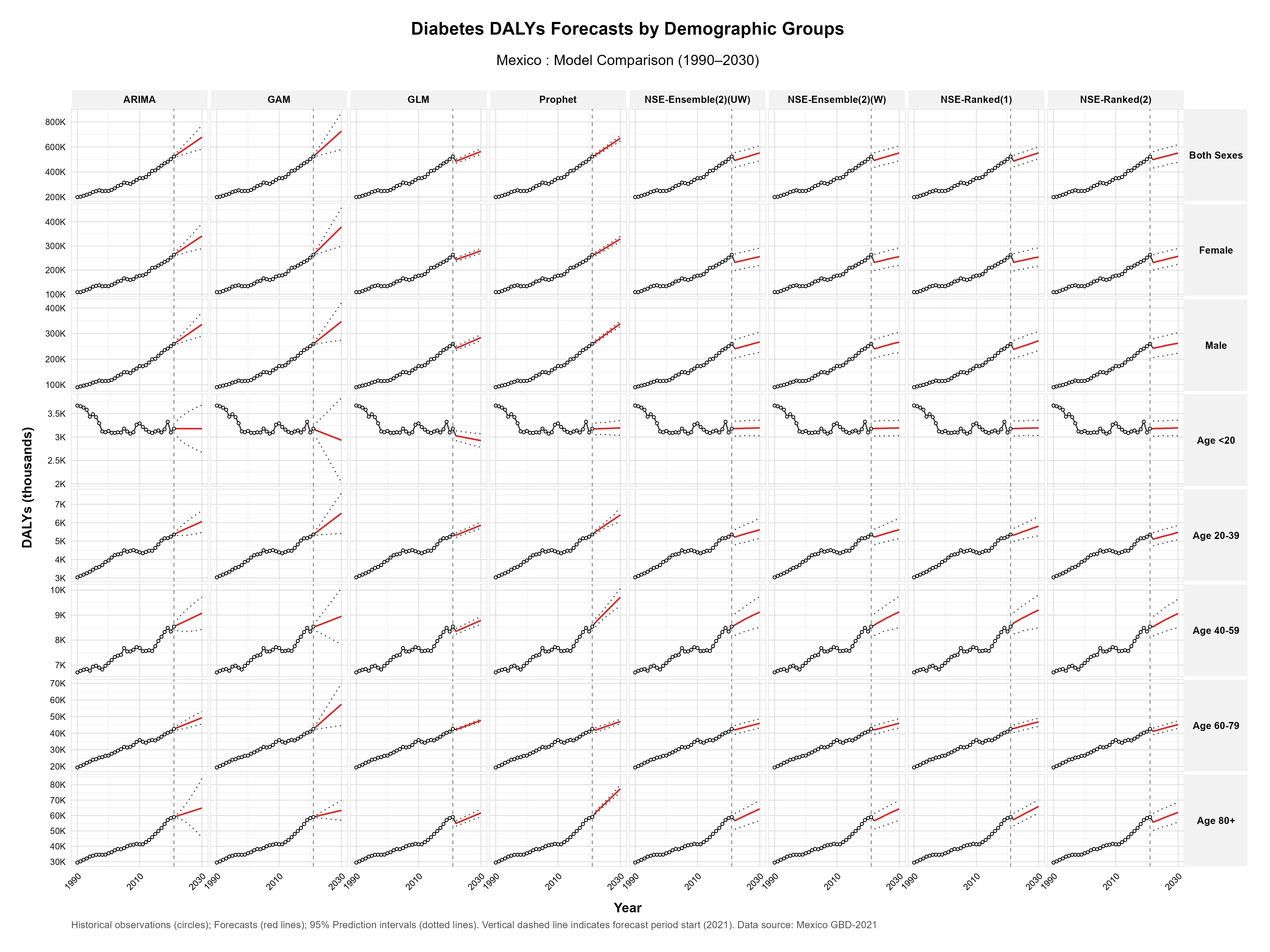

#### Supplementary Fig S5.2 Diabetes DALYs forecasts by demographic groups in USA, 1990-2030.

Model comparison showing historical observations (circles) and forecasts (red lines) with 95% prediction intervals (dotted lines) for diabetes-related disability-adjusted life years across eight forecasting models (ARIMA, GAM, GLM, Prophet, NSE-Ensemble variations, and NSE-Ranked variations). Data are stratified by sex (both sexes, female, male) and age groups (<20, 20-39, 40-59, 60-79, 80+ years). The vertical dashed line indicates forecast period start (2021).

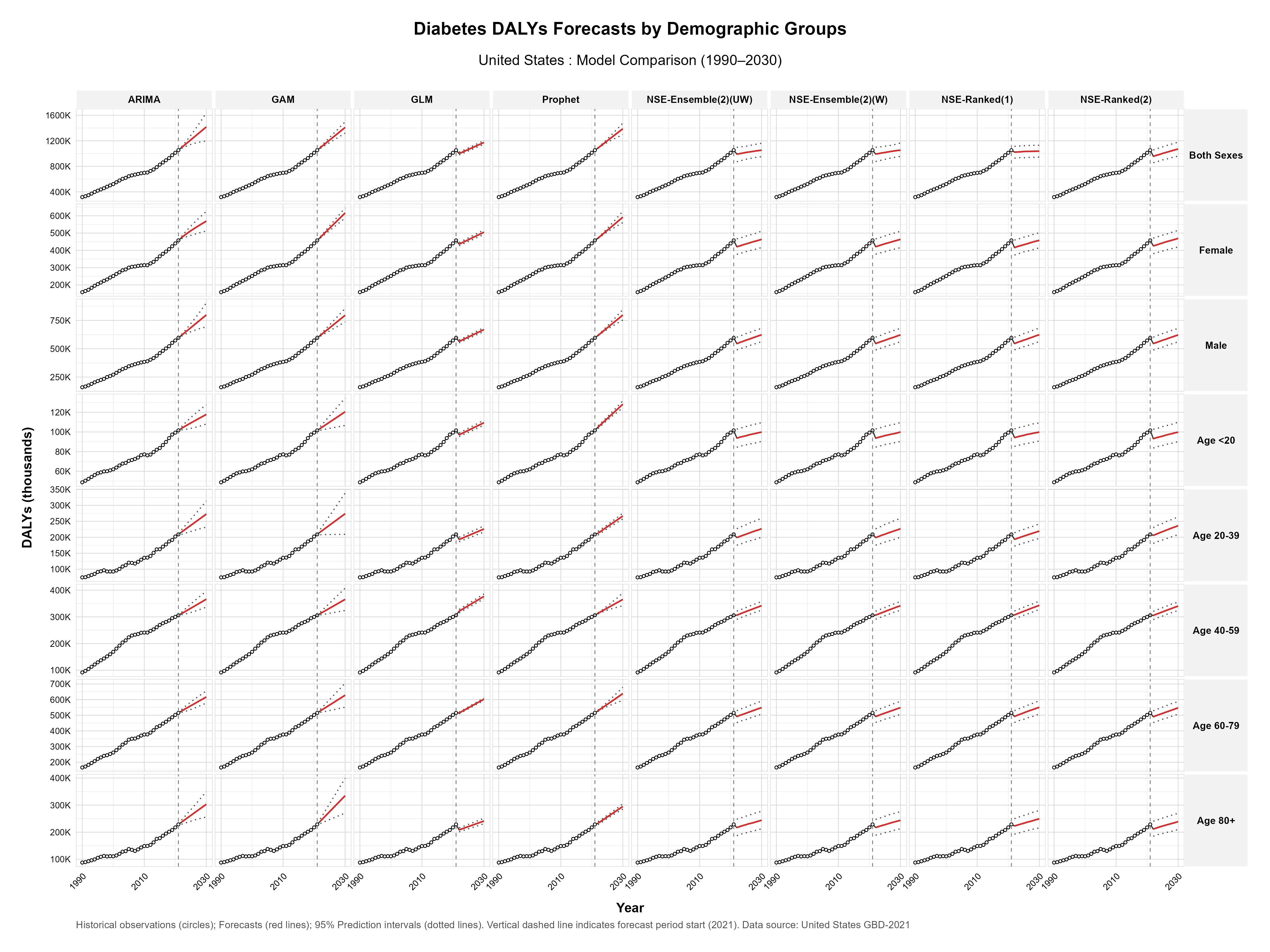
